# Automated Detection of Extrahepatic Bile Duct Stones on Intraoperative Cholangiography Using Deep Learning

**DOI:** 10.64898/2026.08.20.26360965

**Authors:** Yimeng He, Matthew Bloom, Sayeh Mirshojae, Lake Noel, Touseef Qureshi, Yibin Xie, Edward Phillips, Debiao Li, Xiuzhen Huang

**Author notes:** These authors contributed equally to this work (Yimeng He, Matthew Bloom). **Corresponding authors:** Matthew Bloom; Debiao Li; Xiuzhen Huang.

## Abstract

**Objective:** To evaluate the case-level performance of deep-learning segmentation models for detecting extrahepatic bile duct stones on representative intraoperative cholangiography images (IOC) and to characterize the completeness of individual-stone localization.

**Background:** Retained bile duct stones can cause biliary obstruction, cholangitis, and pancreatitis. However false-positive interpretation of filling defects may prompt additional downstream procedures. Computer vision has been applied to biliary anatomy recognition and IOC adequacy assessment, but patient-level stone detection and individual-stone localization remain insufficiently studyed.

**Methods:** Representative IOC images were annotated for extrahepatic biliary anatomy and stones, with case-level stone status established using a composite clinical reference standard. Two deep-learning models were developed to delineate the common bile duct and common hepatic duct and to detect and localize stones. Case-level diagnostic performance was evaluated against the composite clinical reference standard, and individual-stone localization was evaluated against expert-reviewed annotations.

**Results:** On the held-out 125 patients test set, MiT-B2-UNet identified 23 of 25 stone-positive cases and 95 of 100 stone-negative cases, corresponding to a sensitivity of 0.920, specificity of 0.950, and AUC of 0.986. nnU-Net identified 19 of 25 stone-positive cases and 98 of 100 stone-negative cases, corresponding to a sensitivity of 0.760, specificity of 0.980, and AUC of 0.959. At the individual-stone level, MiT-B2-UNet and nnU-Net localized 31 of 59 and 25 of 59 annotated stones, respectively; all annotated stones were localized in 13 of 25 and 12 of 25 stone-positive cases.

**Conclusions:** Deep-learning models can identify stone-positive IOC cases and localize individual stones. This technology may help interpreting the IOCs and reduce retained stones and unnecessary downstream interventions.

## 1 Introduction

Missed common bile duct (CBD) stones during laparoscopic cholecystectomy can lead to adverse events. Clinically significant retained stones occur in approximately 1.8%–2.3% of patients, and approximately one-third present with serious complications such as acute cholangitis^1,2^. The harm of overcalling stones is comparable and more frequently encountered. More than half of patients with abnormal IOCs proceed directly to ERCP, which can itself trigger adverse events such as pancreatitis and, rarely, death^3,4^. Given the large volume of laparoscopic cholecystectomies performed in the United States, the burden of both error directions is substantial^5^. Accurate identification of CBD stones is therefore a high-value clinical target.

Intraoperative cholangiography (IOC) is the most widely used intraoperative imaging technique for laparoscopic cholecystectomy and is endorsed by major surgical guidelines^6,7^. A meta-analysis reported high pooled sensitivity and specificity for stone detection in retrospective settings^8^. However, the reported positive predictive value (PPV) of surgeon-interpreted IOC falls to 48%–63% in real-world studies^9,10^, and inter-surgeon agreement is moderate at best^11,12^.

The suboptimal interpretation performance reflects both task-level difficulty and surgeon-level variability. Filling defects on IOC are visually similar to air bubbles and other artifacts. Surgeon skill is also an important factor; in one cohort, the most experienced surgeons achieved nearly 10-fold higher accuracy than the least experienced^9^. An intraoperative decision-support tool that provides a consistent second read could help surgeons interpret IOC images more accurately and consistently.

Computer vision and deep learning have been increasingly applied to laparoscopic cholecystectomy for intraoperative anatomy recognition and surgical safety assessment^13**?** –15^. For choledocholithiasis specifically, AI has been developed for preoperative imaging^16^ and clinical risk prediction^17–19^, but its application to IOC itself remains largely unexplored. To our knowledge, only one prior study has applied deep learning to IOC interpretation: Badgery et al. trained a multi-class frame-level segmentation model on IOC for autonomous adequacy assessment, with filling defects included as one of several segmentation classes^20^. Case-level AI detection of extrahepatic bile duct stones on IOC has not been previously evaluated. The operating surgeon’s intraoperative decision is made at the case level — whether the cholangiogram contains a filling defect suspicious enough to warrant further imaging, duct exploration, or postoperative ERCP — and a case-level AI tool with high positive predictive value would directly address the dominant interpretive failure of real-time IOC by reducing over-called filling defects while preserving sensitivity to true stones.

The aim of this study was to evaluate whether deep-learning segmentation models can identify cases containing extrahepatic bile duct stones on IOC at the patient level, with stone-level localization assessed as a secondary endpoint.

## 2 Materials and Methods

### 2.1 Study design, cohort, and archived image availability

This retrospective study was approved by the Cedars-Sinai Medical Center Institutional Review Board under protocol STUDY000001017, “AI-aided diagnosis of biliary anatomy,” with waiver of informed consent because the study used de-identified retrospective imaging and clinical data. We reviewed adults 18 years and older who underwent laparoscopic cholecystectomy with IOC at Cedars-Sinai Medical Center between January 1, 2000, and September 1, 2020. Cases were eligible if an archived IOC still image was available and the extrahepatic biliary tree was sufficiently visualized for annotation. Cases were excluded if the IOC was nondiagnostic, the extrahepatic duct could not be adequately visualized, the image file was unavailable or corrupted, or the clinical record did not allow assignment of stone status.

The format of the archived IOC data varied across the study period: some examinations included fluoroscopic video or cine sequences, whereas others contained still images only. To provide a consistent input for analysis, one representative still image was used for each patient. When a sequence or multiple still images were available, trained clinical evaluators selected the frame showing the best visualization of the extrahepatic biliary tree, including the common bile duct and common hepatic duct. Evaluators were blinded to patient identifiers, operative findings, laboratory data, downstream interventions, and clinical follow-up, but had access to the final signed radiology interpretation of the IOC study.

### 2.2 Clinical reference standard

Stone-positive status was assigned using a composite clinical reference standard based on chart review and expert re-review of the IOC studies. Chart review included available radiology reports, operative reports, documentation of intraoperative duct exploration or stone extraction, ERCP findings, and postoperative imaging. Stone-negative status was assigned when there was no documented intraoperative or postoperative evidence of extrahepatic bile duct stones during the index hospitalization and available postoperative follow-up.

### 2.3 Image annotation

A labeling protocol defined key anatomical structures, filling defects, and the cholangiogram catheter; an example is provided in the Supplementary Methods. Ten structure classes plus a background class for unlabeled pixels were defined. The present analysis used the common bile duct, common hepatic duct, and stone labels as model targets. Initial annotations were created by trained clinical annotators using Encord Annotate (Encord, London, United Kingdom) and reviewed by a surgeon experienced in biliary surgery, with disagreements resolved through adjudication. The dataset comprised 1,154 labeled still images from 1,154 unique patients. Images were de-identified, and patients in the held-out test set were distinct from those in the training set to prevent data leakage.

### 2.4 Model development

Images underwent standardized contrast enhancement and anatomy-centered cropping. Two multiclass semantic-segmentation models were developed to identify the common bile duct, common hepatic duct, and stones: the default 2-dimensional nnU-Net framework^21^ and a U-Net decoder^22^ with a pretrained MiT-B2 encoder^23^ and an ROI-conditioned texture-contrast enhancement module. Models were developed using fivefold cross-validation within the training cohort. The five fold-specific models for each architecture were combined by averaging their per-pixel probabilities and evaluated once on the independent held-out test set. Complete preprocessing, architecture, augmentation, and training specifications are provided in the Supplementary Methods.

### 2.5 Outcomes and statistical analysis

The cohort was partitioned at the patient level into a training set (*n* = 1,029) and an independent held-out test set (*n* = 125). Both architectures were developed using fivefold cross-validation on the training set, stratified by stone status. For each architecture, the five fold-specific models were combined by averaging their per-pixel softmax outputs; the per-pixel argmax of the mean output was used as the ensemble segmentation. No fold weighting, calibration, or post-hoc thresholding was applied. All metrics were computed once on the resulting ensemble over the held-out test set, which contained 25 stone-positive and 100 stone-negative cases.

The primary endpoint was case-level identification of stone-positive cases. A case was classified as model-positive if its ensemble segmentation contained at least one pixel assigned to the stone class. The continuous score used for receiver operating characteristic analysis was the maximum stone-channel probability across pixels. Sensitivity (denominator = 25), specificity (denominator = 100), positive predictive value (PPV), AUC, and F1 were reported. Because the test set was enriched for stone-positive cases, PPV reflects the test-set composition rather than the clinical base rate.

Secondary localization endpoints used connected annotated stone components as the unit of analysis. An annotated component was detected if it had any overlap with the predicted stone mask. Stone-level PPV used one-to-one matching under the same any-overlap criterion. Complete per-patient stone detection was defined as localization of every annotated stone in a stone-positive case. Pixel-level duct and stone segmentation were evaluated as technical secondary endpoints and are reported in the Supplementary Results.

Ninety-five percent confidence intervals were obtained using Wilson intervals for binomial proportions, the DeLong method for AUC, stratified percentile bootstrap for F1, and patient-cluster percentile bootstrap for stone-level metrics, as appropriate. Bootstrap analyses used 10,000 replicates. Full ensemble, matching, and interval procedures are provided in the Supplementary Methods (Statistical Procedures subsection). Between-model comparisons were descriptive; the study was not designed for formal superiority testing.

## 3 Results

### 3.1 Cohort characteristics

The final analytic cohort comprised 1,154 IOC studies from 1,154 unique patients, including 200 cases with extrahepatic bile duct stones (positive cases) and 954 cases without (negative cases). Demographic and clinical characteristics are summarized in Table 1. The cohort was partitioned at the patient level into 1,029 training cases and 125 independent test cases. The test set comprised 25 positive and 100 negative cases (20% positive), broadly consistent with the positive-case proportion of the source cohort (17.3%) while preserving sufficient positives for case-level sensitivity estimation.

**Table 1:** Demographic and clinical characteristics of the study cohort.

| Characteristic | Overall (N = 1,154) |
| --- | --- |
| *Sex, n (%)* |  |
| Female | 706 (61.2) |
| Male | 384 (33.3) |
| Missing | 64 (5.5) |
| Age, years, median [IQR] | 52.0 [39.0, 67.0] |
| *Ethnicity, n (%)* |  |
| Hispanic/Latino | 304 (26.3) |
| Non-Hispanic/Latino | 778 (67.4) |
| Missing/other | 72 (6.2) |
| *Race, n (%)* |  |
| White | 765 (66.3) |
| Black | 88 (7.6) |
| Asian | 91 (7.9) |
| Other/unspecified | 146 (12.7) |
| Missing | 64 (5.5) |
| BMI, kg/m<sup>2</sup>, median [IQR] | 28.3 [24.4, 33.0] |

*Values are presented as n (%) for categorical variables and median [interquartile range] for continuous variables. Demographic features were not used as model inputs and are reported for descriptive purposes only. Percentages may not sum to 100 due to rounding. BMI, body mass index; IQR, interquartile range.*

### 3.2 Case-level detection performance

Case-level diagnostic performance on the held-out test set is summarized in Figure 1. MiT-B2-UNet achieved a sensitivity of 0.920 (95% CI 0.750–0.978), specificity of 0.950 (0.888– 0.978), AUC of 0.986 (0.959–0.995), PPV of 0.821 (0.644–0.921), and F1 score of 0.868 (0.775–0.960). nnU-Net achieved a sensitivity of 0.760 (0.566–0.885), specificity of 0.980 (0.930–0.994), AUC of 0.959 (0.879–0.987), PPV of 0.905 (0.711–0.973), and F1 score of 0.826 (0.698–0.936).

**Figure 1:**
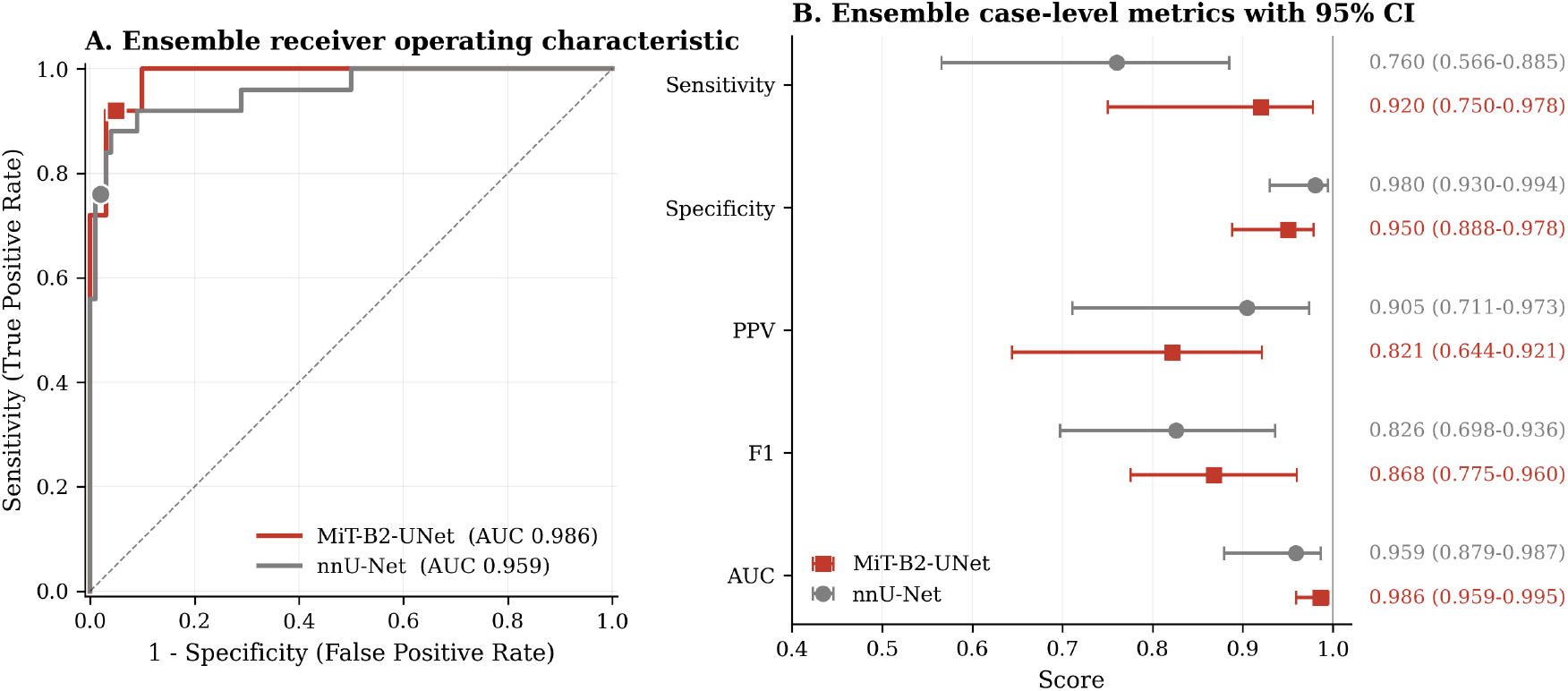
Case-level diagnostic performance on the held-out test set (N = 125; 25 stone-positive and 100 stone-negative cases). (A) Receiver operating characteristic curves for nnU-Net (gray) and MiT-B2-UNet (red), with markers indicating the decision points used for the metrics in panel B. The diagonal dashed line indicates chance performance. (B) Case-level metrics with 95% confidence intervals. AUC, area under the receiver operating characteristic curve; PPV, positive predictive value.

At the prespecified decision rule, the models showed different sensitivity–specificity profiles. MiT-B2-UNet identified 23 of 25 stone-positive cases and 95 of 100 stone-negative cases, whereas nnU-Net identified 19 of 25 stone-positive cases and 98 of 100 stone-negative cases. MiT-B2-UNet therefore had a higher sensitivity point estimate, while nnU-Net had higher specificity and PPV point estimates.

### 3.3 Individual-stone localization

The 25 stone-positive cases in the held-out test set contained 59 annotated stones, with a mean of approximately 2.4 stones per positive case. MiT-B2-UNet localized 31 of 59 stones (52.5%; 95% CI 38.2–71.1), and nnU-Net localized 25 of 59 (42.4%; 28.1–61.5). All annotated stones were localized in 13 of 25 stone-positive cases by MiT-B2-UNet (52.0%; 33.5–70.0) and in 12 of 25 by nnU-Net (48.0%; 30.0–66.5). Thus, both models frequently classified a case correctly after identifying at least one filling defect while failing to localize all stones, particularly in patients with multiple defects (Figure 2).

**Figure 2:**
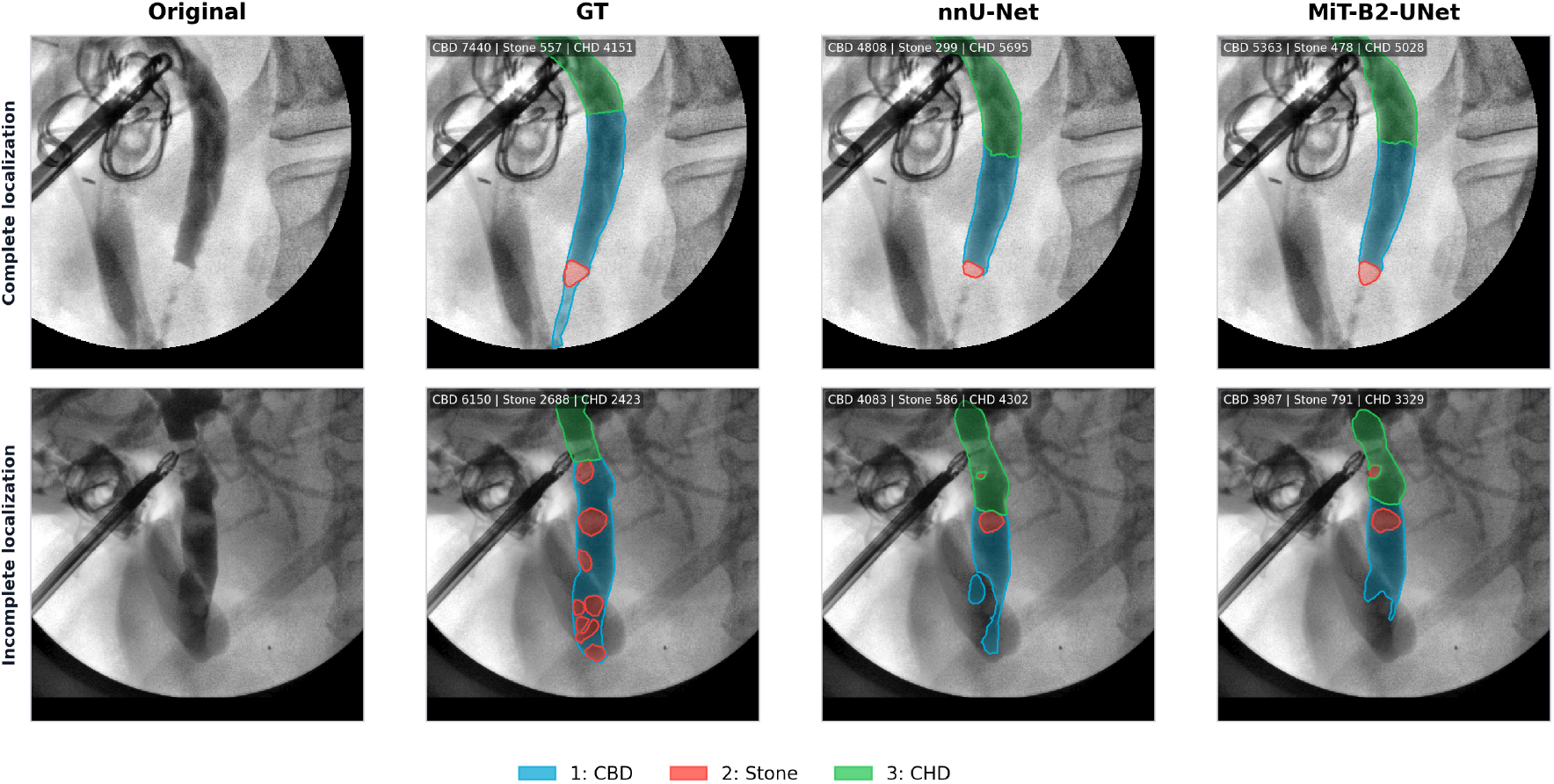
Representative complete and incomplete localization among case-level true-positive predictions. Columns show the original IOC image, ground-truth annotation, nnU-Net prediction, and MiT-B2-UNet prediction. The single-stone case was localized completely by both models, whereas both models classified the multistone case as positive but failed to identify every filling defect. Overlays indicate the common bile duct (blue), stone (red), and common hepatic duct (green).

MiT-B2-UNet produced 36 predicted stone components, 28 of which overlapped an annotated stone (stone-level PPV 77.8%; 62.9–90.5). nnU-Net produced 27 predicted components, 23 of which overlapped an annotated stone (stone-level PPV 85.2%; 66.7–100.0). Among the 100 stone-negative test cases, 5 had at least one false-positive MiT-B2-UNet mark (5.0%; 2.2–11.2), and 2 had at least one false-positive nnU-Net mark (2.0%; 0.6–7.0). Complete results are reported in Table 2.

**Table 2:** Stone-level detection performance.

| Endpoint | nnU-Net | MiT-B2-UNet |
| --- | --- | --- |
| Annotated stones | 59 | 59 |
| Detected stones (of 59 annotated) | 25/59 | 31/59 |
| Stone-level detection rate (%) | 42.4 (28.1–61.5) | 52.5 (38.2–71.1) |
| Per-patient complete stone detection | 12/25 | 13/25 |
| Per-patient complete stone detection rate (%) | 48.0 (30.0–66.5) | 52.0 (33.5–70.0) |
| Predicted stones | 27 | 36 |
| Stone-level PPV (matched/predicted) | 23/27 | 28/36 |
| Stone-level PPV (%) | 85.2 (66.7–100.0) | 77.8 (62.9–90.5) |
| Stone-negative cases with any false-positive mark | 2/100 | 5/100 |
| Stone-negative false-positive case rate (%) | 2.0 (0.6–7.0) | 5.0 (2.2–11.2) |

*Percentages are reported with 95% confidence intervals. Stone-level detection was defined as the proportion of annotated stones with any overlapping predicted stone region. Confidence intervals for stone-level detection and stone-level PPV were estimated using patient-cluster percentile bootstrap resampling with 10,000 replicates. Per-patient complete-detection and stone-negative false-positive confidence intervals used Wilson intervals. Stone-level PPV used one-to-one matching between predicted and annotated stones under an any-overlap criterion.*

### 3.4 Clinically categorized error modes

Both models also delineated the extrahepatic biliary anatomy; complete duct and stone segmentation metrics are reported in Supplementary Table S1.

Two stone-positive cases were missed by both ensembles. One contained a small stone near the 8.6th cohort percentile of stone size (Figure 3A). The other contained a stone near the cohort median (approximately the 55th percentile) that abutted a low-contrast duct segment and lacked the sharp filling-defect boundary seen in more conspicuous stones (Figure 3B). These examples indicate that shared false negatives were associated with both small size and visual subtlety.

**Figure 3:**
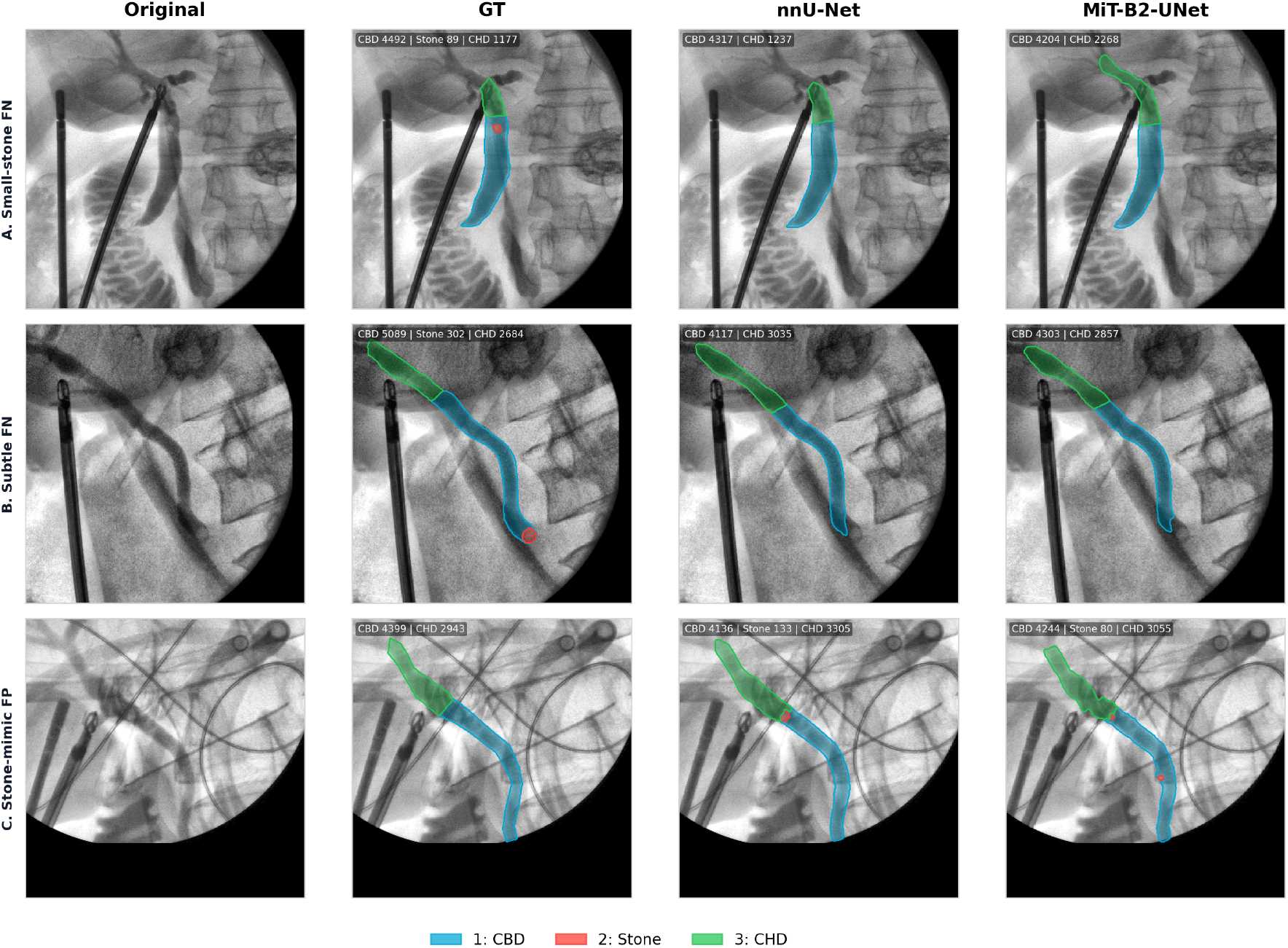
Representative error modes shared by both models on the held-out test set. Columns show the original IOC image, ground-truth annotation, nnU-Net prediction, and MiT-B2-UNet prediction. (A) Small-stone false negative. (B) Visually subtle false negative.(C) Stone-mimic false positive. Overlays indicate the common bile duct (blue), stone or filling defect (red), and common hepatic duct (green).

One stone-negative case was marked positive by both ensembles (Figure 3C). The predicted region was near the distal common bile duct and visually resembled a filling defect, illustrating how stone-mimicking intraductal appearances may produce false-positive predic-tions. These examples characterize technical failure modes on archived still images; they do not establish how the models would perform with the temporal information available during live IOC interpretation.

## 4 Discussion

Artificial intelligence has moved rapidly from research benchmarks into clinical workflow, and surgery is no exception. Recent advances in computational power and more sophisticated algorithms have made it increasingly viable to deploy AI in the operating room (OR). Prior work such as automatic Critical View of Safety (CVS) assessment has explored applying AI at different stages of laparoscopic cholecystectomy and has shown promising results^13–15,24**?**^. To our knowledge, this is the first case-level evaluation of AI for stone detection on IOC, laying the groundwork for real-time AI interpretation in the operating room.

On the held-out test set, both models demonstrated high case-level discrimination for stone-positive IOC cases, with AUCs of 0.959–0.986, sensitivities of 0.760–0.920, and specificities of 0.950–0.980. Individual-stone localization was less complete: the models detected 42.4%–52.5% of annotated stones and localized all stones in 48.0%–52.0% of stone-positive cases. Accordingly, correct case-level classification often reflected detection of at least one filling defect rather than complete representation of the annotated stone burden. Both models also delineated the CBD and CHD, providing anatomic context for the predicted filling defects. The principal clinical finding is therefore the distinction between identifying a stone-positive IOC case and completely localizing every stone.

Cholangiogram interpretation is not a simple task even for experienced surgeons. Nor-mally the IOC images are interpreted in a multi-frame setting: surgeons watch contrast move through the duct and observe whether suspected filling defects persist or disappear after flushing. Without the information from other frames, it is much harder to distinguish real stones from air bubbles or other contrast-streaming artifacts. In addition, the task is more difficult when multiple filling defects overlap or partially obscure one another, and surgeons themselves often disagree on whether subtle defects represent true stones^11,12^.

From a surgical perspective, the results show promise for use as an intraoperative second reader: a system that flags a potentially abnormal cholangiogram and prompts the surgeon to review the image more carefully. This is especially valuable for reducing overcalls. Under intraoperative pressure, surgeons tend to flag most suspicious defects as stones^9,10^. When the surgeon suspects a stone but the model does not, the model result may prompt a second review and increase confidence that the apparent filling defect is not a true stone, potentially reducing unnecessary ERCPs^3,4^. For surgical trainees, the model can provide a consistent starting point for IOC interpretation. Its standardized output may help reduce inter-reader variability — a meaningful benefit given documented skill deficiencies among residents^25^.

There are limitations. First, this was a single-center retrospective study spanning approximately 20 years, during which fluoroscopy systems, contrast protocols, and image acquisition practices have evolved; performance may not generalize to other institutions or to current imaging standards. Second, for each case the analysis used a single representative IOC frame selected by trained clinical evaluators for clarity; model performance is likely optimistic compared to deployment on all frames. Third, the held-out test set included only 25 stone-positive cases, resulting in wide confidence intervals, particularly for stone-level and complete per-patient metrics. Fourth, the segmentation pipeline relied on a predefined CBD region of interest, and errors in CBD localization could propagate to the stone-detection stage. Finally, positive predictive value will vary with stone prevalence across clinical settings.

Looking ahead, further studies will follow to validate the system in the OR, define the optimal interface for intraoperative feedback, assess its value as a second reader during IOC interpretation, and, ultimately, reduce both retained stones and unnecessary ERCPs.

## 5 Conclusions

This study suggests that AI systems can be used to distinguish stone-positive from stone-negative patients on IOC images with a high degree of performance. As additional evidence emerges on the safety and effectiveness of AI in the OR, such tools have the potential to serve as an intraoperative second reader and eventually reduce both retained stones and unnecessary ERCPs.

## Data Availability

The data that support the findings of this study are not publicly available due to patient privacy and institutional restrictions but may be available from the corresponding author on reasonable request and with appropriate institutional approvals.

