## supplemental materials for "Automated Detection of Extrahepatic Bile Duct Stones on Intraoperative Cholangiography Using Deep Learning"

### S1 Supplementary Methods

#### S1.1 Image annotation protocol

The complete annotation ontology comprised 10 structure classes plus a background class. Figure S1 provides an example of the annotation labels.

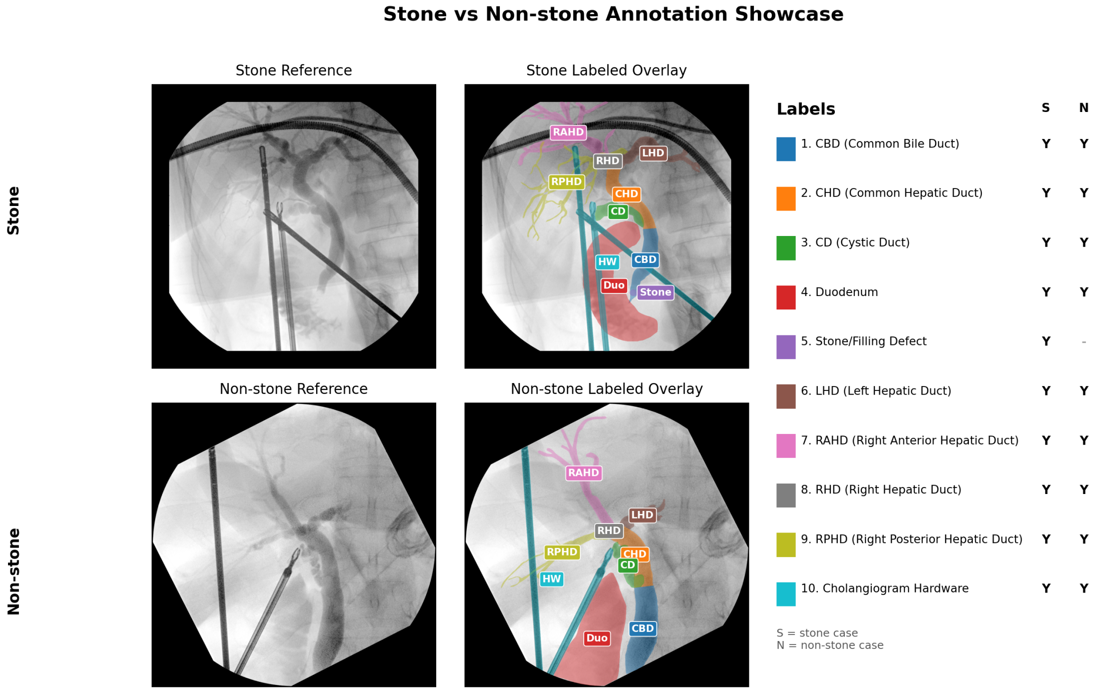


Figure S1: Example of the complete 10-class annotation protocol.

#### S1.2 Image preprocessing

##### Resizing

Each raw IOC still image was resized to $512 \times 512$ pixels using bilinear interpolation (`cv2.resize` with `INTER_LINEAR`). Pixel intensities were retained in their original 8-bit unsigned range before histogram equalization.

##### Collimator-border detection

A binary mask of the imaged fluoroscopic field was produced by thresholding the resized image at an intensity of 20% to separate the low-intensity collimator ring from the contrast-bearing diagnostic region. The largest connected foreground component was retained as the collimator-interior mask.

##### Border-aware CLAHE

Contrast-limited adaptive histogram equalization[1] was performed using OpenCV[2] (`cv2.createCLAHE(clipLimit=3.0, tileGridSize=(8,8))`) and applied to the entire $512 \times 512$ image. The collimator-interior mask was then used to restore the original pixel values outside the collimated region so that histogram-equalization statistics inside the imaged field were not contaminated by the dark collimator ring.

##### Anatomy-centered cropping

A $384 \times 384$ region centered on the junction of the common bile duct (CBD) and common hepatic duct (CHD) was extracted from the contrast-equalized $512 \times 512$ image. Let $\mathcal{M}_{\mathrm{CBD}}$ and $\mathcal{M}_{\mathrm{CHD}}$ denote the binary masks of the two ducts. The crop center $(x_c, y_c)$ was defined as the centroid of $\mathcal{M}_{\mathrm{CBD}} \cup \mathcal{M}_{\mathrm{CHD}}$. The crop window $[x_c - 192,\ x_c + 192] \times [y_c - 192,\ y_c + 192]$ was extracted. If the window extended beyond the image bounds, the image was zero-padded so that the network input was always $384 \times 384$.

##### Two-pass inference

During training, the manually annotated CBD and CHD masks were used to compute the crop center. At inference, the segmentation network was applied in two passes. The first pass produced provisional CBD and CHD masks; those masks were used to define the anatomy-centered crop and ROI prior for the second pass.

#### S1.3 Model architectures

##### nnU-Net

nnU-Net[3] was used in its default 2-dimensional configuration. The framework's automatic experiment planner determined the patch size, network depth, number of feature channels per stage, normalization scheme, downsampling factors, and per-fold training schedule from the dataset fingerprint. No manual modifications were made to the nnU-Net pipeline beyond the data partition and inclusion of the CBD, CHD, and stone foreground classes.

##### MiT-B2-UNet

MiT-B2-UNet was implemented with the `segmentation_models_pytorch` library[4] (`smp.Unet`). The Mix-Transformer MiT-B2 encoder[5] had four hierarchical stages that produced feature maps at strides $\{4, 8, 16, 32\}$ relative to the $384 \times 384$ input. Encoder weights were initialized from ImageNet-1K classification pretraining; the U-Net decoder was randomly initialized. The decoder used the four encoder feature maps as skip connections.

##### ROI-conditioned texture-contrast enhancement module

To amplify subtle stone cues, the module computed an additive enhancement payload that combined a Laplacian high-pass texture response and an ROI-conditioned contrast response.

A texture feature $t_s$ was computed using a high-frequency transform:

$$t_s = \mathrm{HighPass}(f_s), \tag{1}$$

and a contrast feature $c_s$ was computed using a convolutional block conditioned on the ROI prior:

$$c_s = \psi_s([f_s, R_s]). \tag{2}$$

The two signals were averaged, and their magnitude was bounded using tanh:

$$p_s = \tanh\left(\tfrac{1}{2}(t_s + c_s)\right). \tag{3}$$

The payload was then injected into the ROI-reweighted features under the ROI gate:

$$f'_s = \tilde{f}_s + \lambda_s\, g_s \odot p_s, \tag{4}$$

where $\lambda_s$ was a learnable per-scale strength parameter. The update was applied at multiple encoder scales $\mathcal{S}$ (for example, $s \in \{3, 4, 5\}$), and the refined features $\{f'_s\}$ were passed to the decoder to produce the final prediction.

#### S1.4 Training

##### Data augmentation

For MiT-B2-UNet, training-time augmentation was implemented with the `albumentations` library[6] and applied synchronously to the input image and the CBD, CHD, and stone masks. The pipeline contained the following independently sampled transforms:

- `HorizontalFlip` ($p = 0.5$);
- `VerticalFlip` ($p = 0.5$);

- `Rotate` with an angle sampled uniformly from $[-30°, +30°]$ ($p = 0.5$);
- `ShiftScaleRotate` with translation up to 10% of the image dimensions in each direction and isotropic scaling within ±10%, with no additional rotation (`rotate_limit` $= 0$, $p = 0.5$);
- `GridDistortion` with a $5 \times 5$ control grid and per-cell distortion limit of 0.3 ($p = 0.3$);
- `ElasticTransform` with displacement magnitude $\alpha = 1$, Gaussian smoothing $\sigma = 50$, and affine displacement $\alpha_{\text{affine}} = 50$ ($p = 0.2$);
- `RandomBrightnessContrast` with symmetric brightness and contrast limits of ±0.2 ($p = 0.4$);
- `GaussNoise` with per-pixel variance sampled uniformly from $[0.001, 0.01]$ in normalized pixel-intensity units ($p = 0.2$).

Geometric transforms used bilinear interpolation for the image and nearest-neighbor interpolation for the masks so that label identities were preserved.

#### Training protocol

The nnU-Net architecture was trained with the framework's default 2-dimensional configuration. Its automatic experiment planner determined the patch size, network depth, normalization scheme, optimizer, learning-rate schedule, and per-fold training duration from the dataset fingerprint; no manual hyperparameter modifications were applied.

MiT-B2-UNet was trained with a combined Dice and cross-entropy loss and the AdamW optimizer[7] at a peak learning rate of $1 \times 10^{-4}$ and batch size 16, using a cosine annealing learning-rate schedule[8] with a 100-epoch linear warm-up. Training was run for a maximum of 1,000 epochs with early stopping on validation loss (patience, 75 epochs). Data augmentation was performed with `albumentations`[6]. Training was performed on a single NVIDIA L40S GPU.

#### S1.5 Statistical procedures

**Ensemble construction.** For each architecture, the five cross-validation-fold models were combined by taking the pixel-wise arithmetic mean of their softmax outputs. Let $\mathbf{s}^{(k)}(x, y, c)$ denote the probability assigned to class $c$ at pixel $(x, y)$ by the fold-$k$ model $(k = 1, \ldots, 5)$. The ensemble tensor was $\bar{\mathbf{s}}(x, y, c) = \frac{1}{5}\sum_{k=1}^{5} \mathbf{s}^{(k)}(x, y, c)$, and the per-pixel argmax over classes was used as the ensemble segmentation. No fold weighting, calibration, or post-hoc thresholding was applied. All metrics were computed once on this ensemble over the held-out test set ($n = 125$; 25 stone-positive and 100 stone-negative cases).

**Confidence-interval framework.** Confidence-interval methods were matched to the variance structure of each metric: Wilson 95% intervals for binomial proportions, DeLong 95% intervals for AUC, and percentile bootstrap intervals with 10,000 resamples for metrics without a closed-form variance estimator. These intervals quantify sampling uncertainty in the held-out test set conditional on the trained ensemble and do not incorporate uncertainty from model training or annotation variability.

**Case-level metrics.** A case was classified as model-positive if the ensemble segmentation contained at least one pixel assigned to the stone class. The continuous score used for AUC and receiver operating characteristic analysis was the maximum stone-channel probability across pixels. Sensitivity (TP/(TP+FN); denominator = 25), specificity (TN/(TN+FP); denominator = 100), and PPV (TP/(TP+FP)) were reported with Wilson 95% confidence intervals. AUC was treated as a U-statistic over pairwise comparisons of positive and negative case scores and reported with DeLong 95% confidence intervals. Because AUC approached the upper boundary, intervals were computed on the logit scale and back-transformed by the inverse logit. F1 was reported with stratified percentile bootstrap 95% confidence intervals. In each of 10,000 replicates, the 25 stone-positive and 100 stone-negative cases were resampled with replacement separately to preserve class balance. Because the design prevalence was 20%, PPV reflects the test-set composition and should not be interpreted as the value expected at the clinical base rate.

**Segmentation metrics.** The Dice similarity coefficient was calculated separately for each held-out case and summarized as the mean per-case value. Ninety-five percent confidence intervals were estimated by nonparametric case-level bootstrap with 10,000 replicates. CBD and CHD Dice were calculated among cases in which the corresponding structure was annotated. Stone Dice was calculated among stone-positive cases only; stone-positive cases without a predicted stone mask were assigned a Dice of 0, and stone-negative cases were excluded.

**Stone-level metrics.** Connected annotated stone components were the unit of analysis. An annotated component was detected if it had any overlap with the predicted stone mask. Because multiple components could occur within the same patient, 95% confidence intervals for the detection rate were estimated using patient-cluster nonparametric bootstrap with 10,000 replicates. In each replicate, stone-positive patients were sampled with replacement, all annotated components from selected patients were retained, and the detection rate was recalculated. Stone-level PPV was the number of predicted components matched to annotated components divided by the total number of predicted components. Matching was one-to-one under an any-overlap criterion, and its confidence intervals were estimated by patient-cluster bootstrap over all test patients. Complete detection of all annotated stones within a stone-positive case and the proportion of stone-negative cases with any false-positive stone mark were reported with Wilson 95% confidence intervals.

### S2 Supplementary Results

#### S2.1 Segmentation performance

Pixel-level segmentation performance on the held-out test set is summarized in Table S1. For the common bile duct ($n = 124$), nnU-Net and MiT-B2-UNet achieved mean Dice scores of 0.854 (95% CI 0.826–0.878) and 0.853 (0.824–0.879), respectively. For the common hepatic duct ($n = 122$), the corresponding mean Dice scores were 0.784 (0.752–0.813) and 0.767 (0.731–0.800). Stone Dice, restricted to the 25 stone-positive cases, was 0.448 (0.310–0.587) for nnU-Net and 0.460 (0.336–0.581) for MiT-B2-UNet.

Table S1: Segmentation performance on the held-out test set.

| Class | Metric | nnU-Net | MiT-B2-UNet |
|---|---|---|---|
| CBD ($n = 124$) | Dice | **0.854 (0.826–0.878)** | 0.853 (0.824–0.879) |
| CHD ($n = 122$) | Dice | **0.784 (0.752–0.813)** | 0.767 (0.731–0.800) |
| **Stone*** ($n = 25$) | Dice | 0.448 (0.310–0.587) | **0.460 (0.336–0.581)** |

*Values are mean per-case metrics with 95% confidence intervals estimated by nonparametric bootstrap resampling of held-out cases with 10,000 replicates. Common bile duct and common hepatic duct metrics were calculated among cases in which the structure was annotated. *Stone segmentation metrics were calculated among stone-positive cases only.*

#### S2.2 Architecturally divergent cases

The case-level differences between the two models arose from a small set of borderline cases on which their ensemble predictions differed. MiT-B2-UNet identified four stone-positive cases that nnU-Net missed, whereas nnU-Net correctly classified four stone-negative cases that MiT-B2-UNet marked positive. These eight of 125 cases account for most of the observed difference in the case-level point estimates. This analysis is descriptive and does not establish that architectural features caused the observed differences.

[7] Loshchilov Ilya, Hutter Frank. Decoupled weight decay regularization *arXiv preprint arXiv:1711.05101.* 2017.

[8] Loshchilov Ilya, Hutter Frank. Sgdr: Stochastic gradient descent with warm restarts *arXiv preprint arXiv:1608.03983.* 2016.
